# Automated machine learning of echocardiographic strain enables identification of early myocardial changes in pre-symptomatic *TTR* carriers

**DOI:** 10.64898/2026.03.04.26347545

**Authors:** Amit Weigman, Wenli Zhao, Steve L. Liao, Maria Giovanna Trivieri, Samuel Madiman, Stamatios Lerakis, Eimear E. Kenny, Noura S. Abul-Husn, Vikas Pejaver, Amy R. Kontorovich

## Abstract

**Objectives:** To identify unique echocardiographic signatures associated with *TTR*+ carrier status preceding onset of cardiac amyloidosis.

**Background:** Carrier status for the most common pathogenic *TTR* variant in the United States, Val142Ile (V142I), found in 4% of African Americans (AA) and 1% of Hispanic/Latino (H/L) individuals, confers a 40-60% lifetime risk of developing variant transthyretin amyloidosis (ATTRv), including cardiac amyloidosis (CA) and heart failure (HF). Myocardial amyloid deposition is believed to progress over many years. Genomic screening programs and familial cascade genetic testing are increasingly uncovering pre-symptomatic *TTR*+ carriers, yet no guidelines exist to pragmatically risk stratify these individuals for CA.

**Methods:** V142I+ carriers (cases) without prior diagnoses of amyloidosis or HF were identified among Bio*Me* biobank participants with available exome sequencing data linked to electronic health records (EHRs) including at least one available echocardiogram. Controls were biobank participants with normal *TTR* sequencing who were age-, sex- and ancestry-matched to cases. Speckle-tracking echocardiography (STE) was applied to images and conventional and strain measurements were evaluated by univariate analyses. A random forest model was trained using a minimal redundancy maximal relevance (mRMR, applied to mitigate overfitting) feature set and evaluated by 5-fold cross-validation to minimize optimism bias. Discriminatory performance was assessed using the area under the receiver operating characteristic curve (AUC).

**Results:** 49 *TTR*+ (100% V142I, median age 61 years, 69.4% female) and 45 matched *TTR*-biobank participants were included in the model development cohort. STE generated approximately 200 features. Univariate analyses revealed no significant differences between carriers and controls on any individual strain or conventional echocardiographic measurements including global longitudinal, right ventricular and left atrial strain. mRMR feature selection resulted in a set of 15 features retained for all downstream modeling, integrating global amyloid signatures, regional inferolateral strain abnormalities, layer-specific deformation, and mechanical timing heterogeneity. Using this feature set, the model achieved good discrimination (AUC=0.76). Feature importance analysis highlighted relative apical sparing, inferolateral strain reduction, and basal-apical timing gradients as key contributors to model performance. External validation (n=115) confirmed good model discrimination (AUC=0.781, 95% CI: 0.688-0.869, sensitivity 0.983).

**Conclusions:** Machine learning applied to routinely acquired echocardiographic data can identify subtle myocardial abnormalities associated with *TTR* V142I carrier status prior to development of CA. Key model features are physiologically relevant to known echocardiographic characteristics of overt CA. Genotype-guided echocardiographic surveillance may be a scalable strategy for early detection of CA risk.

## Introduction

Transthyretin amyloidosis (ATTR) is a progressive and debilitating disease caused by either pathogenic variants in the *TTR* gene (variant ATTR, ATTRv) or an age-related process (wild-type ATTR, ATTRwt), leading to cardiac amyloidosis (CA) and heart failure (HF), and/or polyneuropathy (PN)(1). Over 130 *TTR* variants have been identified as causative for ATTRv(2) but the most clinically relevant in the United States is V142I; this is likely the most common single dominant pathogenic variant worldwide—recent studies estimate prevalence at 1:330 individuals(3)—and it is enriched in African American (AA) (1:25)(4) and Hispanic/Latino (H/L) (1:100)(5) populations. The V142I variant is estimated to have a 40-60% lifetime penetrance for ATTRv-associated HF and PN(5–7), with higher penetrance in individuals ≥ age 60. In fact, V142I alone attributes 27% risk for incident HF, with traditional risk factors (hypertension, diabetes, coronary artery disease) augmenting the cumulative risk of HF(8). Largely due to under-detection, a recent health burden analysis concluded that V142I contributes to 1 million years of life lost among US Black individuals ≥ age 50 that go without ATTRv treatments(9).

In most series, >90% of patients undergoing evaluation of ATTR-CA (typically prompted by clinical recognition of signs or symptoms) already have Class II or higher HF(10), indicating advanced disease. FDA-approved TTR stabilizer(11,12) and silencer(13,14) therapies have been shown to improve symptoms, functional endpoints and biomarkers of ATTRv but do not remove amyloid deposits from tissues nor reduce mortality in patients with advanced disease. In the ATTR-ACT trial comparing efficacy of the TTR tetramer stabilizer tafamidis against placebo, the overall positive outcome of reduction in all-cause mortality and cardiovascular-related hospitalizations was driven by the response in the least symptomatic group of CA patients (NYHA Class I-II)(15). More recent trials have enrolled a less symptomatic subset of patients with ATTRv, resulting in even stronger treatment effects; >85% of patients in the HELIOS-B trial in which vutrisiran had a hazard ratio of 0.72 for risk of death from any cause and recurrent cardiovascular events were classified as NYHA class I or II(16). Furthermore, a long term extension study of the ATTR-ACT population found that patients treated with tafamidis had significantly better survival over those first treated with placebo and then switched to tafamidis after 30 months, supporting that early treatment improves outcomes(17). These observations have led to calls for earlier screening and detection(18) and use of TTR stabilizing therapies in *TTR*+ carriers as early as possible(19). Expanded access to clinical genetic testing, genomic screening programs, and familial cascade testing has led to the identification of growing numbers of pre-symptomatic *TTR*+ individuals. Tools to identify early features of CA in such *TTR*+ carriers are therefore urgently needed and may eventually enable earlier diagnosis of ATTRv and improved outcomes.

Although CA is typically recognized only after symptoms emerge and, in V142I disease, often after the 6^th^ decade of life, we hypothesized that amyloid deposition commences earlier and could be gleaned through unique machine learning (ML)-derived echocardiographic signatures. Using a “genotype-first” approach, we retrospectively performed deep learning on echocardiographic data from *TTR*+ and *TTR-* participants in our diverse institutional biobank, Bio*Me*, to identify echocardiographic signatures associated with increased genetic risk for CA.

## Methods

### Study population: development cohort

The study was approved by Mount Sinai’s Institutional Review Board (GCO # 19-01925 ISMMS). Participants were identified through their participation in the electronic health record (EHR)-linked Bio*Me* biobank at the Icahn School of Medicine at Mount Sinai in New York City. Participant enrollment into Bio*Me* was non-selective and occurs primarily through ambulatory care practices across the Mount Sinai Health System. Exome sequencing was available for 30,223 adult Bio*Me* participants through a prior agreement with the Regeneron Genetics Center. We identified participants harboring the *TTR* V142I variant (*TTR*+) and with ≥ 1 echocardiogram suitable for retrospective speckle tracking echocardiography (STE) analysis in their EHR. Controls were Bio*Me* participants without any pathogenic or likely pathogenic variants in *TTR* (*TTR*-) having ≥ 1 echocardiogram suitable for STE in their EHR and were matched to *TTR*+ subjects by sex, self-reported ancestry and age at time of echocardiogram (+/- 1 year). Echocardiograms were screened to determine if images were suitable (i.e. completeness of studies, compatibility of software, quality of images) for post-processing by STE. Participants with any International Classification of Diseases (ICD)-9 or ICD-10 diagnosis codes related to ATTR, HF or cardiomyopathy were excluded.

### Study population: validation cohort

The study was approved by Mount Sinai’s Institutional Review Board (GCO # 19-00565 ISMMS). Cases were drawn from a convenience sample of asymptomatic *TTR*+ carriers identified through their enrollment in a deep cardiac phenotyping study (NIH R01HL155356). Controls were matched from *TTR*-BioMe participants using the same criteria and process described for the development cohort.

### Echocardiographic assessments

Archived transthoracic echocardiograms were analyzed using STE software (TomTec, Philips) in a blinded fashion. Standard apical and short-axis views were selected based on image quality. GLS, segmental longitudinal strain across basal, mid-ventricular, and apical segments, circumferential strain, and right ventricular strain parameters were quantified where feasible.

Subjects with inadequate images for myocardial deformation analysis (defined as inadequate tracking in ≥2 segments) were excluded from downstream analysis. Clinical covariates, qualitative and quantitative echocardiographic parameters were extracted from clinical imaging reports where available.

### Statistical analyses

Univariate comparison of baseline demographic and echocardiographic characteristics between *TTR*+ and *TTR*-groups was performed using the Mann-Whitney U test.

### Feature selection for machine learning

Minimal redundancy maximal relevance (mRMR) feature selection was used to select features that maximize relevance to the outcome (*TTR*+ status) while minimizing redundancy among selected features. This approach is particularly well suited to echocardiographic strain data, where adjacent segments, myocardial layers, and temporally related measurements often convey overlapping information. By penalizing redundancy, mRMR yields a parsimonious feature set that preserves complementary physiologic signals.

### Machine learning model and evaluation

A random forest (RF) classifier was trained using the mRMR-selected feature set. Model evaluation was performed using cross-validation to minimize optimism bias. Discriminatory performance was assessed using the area under the receiver operating characteristic curve (AUC). Feature importance was examined to facilitate physiologic interpretation and to confirm that model behavior aligned with known and hypothesized patterns of amyloid involvement. The final model was then tested in the external validation cohort, with discriminatory performance reported by AUC.

## Results

### Demographics and clinical characteristics of development cohort

49 *TTR*+ cases (ages 24-90 years, 100% V142I) and 45 *TTR*-matched controls (ages 23-90 years) met the inclusion criteria. All subjects were of self-reported AA or H/L ancestry. Baseline characteristics between groups were similar (**Table 1**). There were 25 (51%) *TTR*+ and 27 (60%) *TTR*-subjects ≥ age 60 (p = 0.41), and demographic features were similar between these sub-cohorts.

**Table 1.**
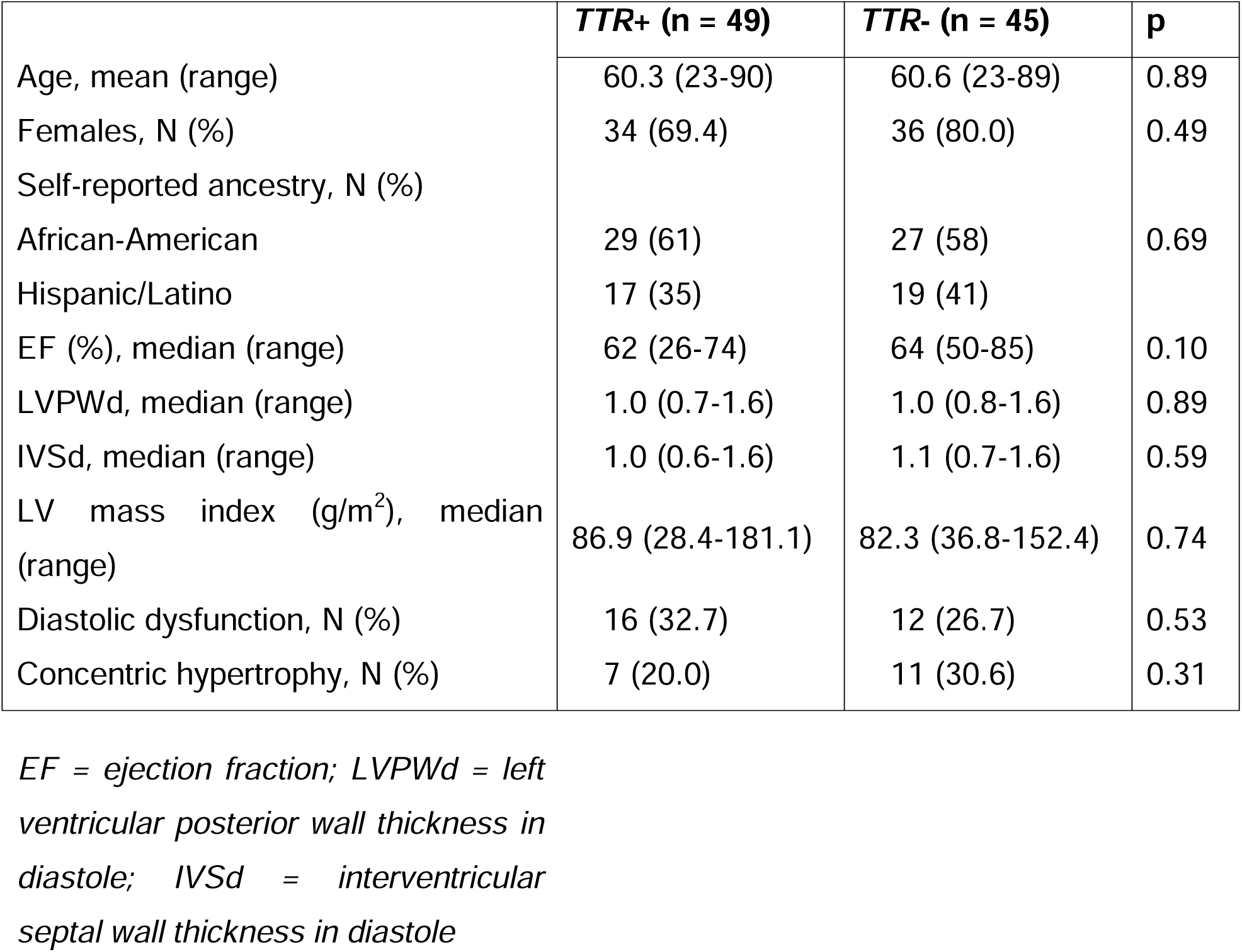
Demographic and standard echocardiographic features of *TTR*+ and *TTR*-participants in the development cohort.

|  | ***TTR*+ (n = 49)** | ***TTR*- (n = 45)** | **p** |
| --- | --- | --- | --- |
| Age, mean (range) | 60.3 (23-90) | 60.6 (23-89) | 0.89 |
| Females, N (%) | 34 (69.4) | 36 (80.0) | 0.49 |
| Self-reported ancestry, N (%) |  |  |  |
| African-American | 29 (61) | 27 (58) | 0.69 |
| Hispanic/Latino | 17 (35) | 19 (41) |  |
| EF (%), median (range) | 62 (26-74) | 64 (50-85) | 0.10 |
| LVPWd, median (range) | 1.0 (0.7-1.6) | 1.0 (0.8-1.6) | 0.89 |
| IVSd, median (range) | 1.0 (0.6-1.6) | 1.1 (0.7-1.6) | 0.59 |
| LV mass index (g/m<sup>2</sup>), median (range) | 86.9 (28.4-181.1) | 82.3 (36.8-152.4) | 0.74 |
| Diastolic dysfunction, N (%) | 16 (32.7) | 12 (26.7) | 0.53 |
| Concentric hypertrophy, N (%) | 7 (20.0) | 11 (30.6) | 0.31 |

*EF* = ejection fraction; *LVPWd* = left ventricular posterior wall thickness in diastole; *IVSd* = interventricular septal wall thickness in diastole

### Echocardiographic features

In addition to conventional echocardiographic measures (e.g., ejection fraction, wall thickness), automated parsing and post-processing of STE data generated approximately 200 candidate features. These included raw strain values at end-systole and peak contraction, layer-specific (endocardial and mid-myocardial) measures, regional gradients (e.g., apical-to-basal ratios), dispersion metrics, and time-to-peak strain features capturing mechanical timing and dyssynchrony. Engineered features reflecting known amyloid physiology, such as relative apical sparing indices(20), were also included.

### Univariate analysis

Consistent with prior univariate analyses, individual global and segmental strain parameters did not reliably differentiate *TTR*+ carriers from *TTR*-controls. Global longitudinal, right ventricular and left atrial strain measures, as well as qualitative or quantitative apical sparing indices showed substantial overlap between groups (**Table 2**).

**Table 2.** Univariate Comparison of Speckle Tracking Echocardiography-Derived Strain Measurements Between Groups.

|  | ***TTR+* (n = 49)** | ***TTR-* (n = 45)** | **p** |
| --- | --- | --- | --- |
| GLS (%) | -16.8 | -16.8 | 0.95 |
| Basal average strain | 15.9 | 17.3 | 0.33 |
| Mid average strain | 15.9 | 16.1 | 0.87 |
| Apical average strain | 19.4 | 17.0 | 0.15 |
| Segmental SD | 6.1 | 6.3 | 0.95 |

*GLS = global longitudinal strain*

### Machine learning modeling

Initial ML models trained on the full feature set demonstrated clear overfitting, consistent with the high dimensionality of the data relative to sample size and the strong correlations among strain features. To mitigate this, we applied mRMR, which resulted in a reduced set of 15 features integrating global amyloid signatures (e.g., relative apical sparing), regional inferolateral strain abnormalities, layer-specific deformation, and mechanical timing heterogeneity (**Figure 1**). These features were retained for all downstream modeling. When multivariate ML modeling was applied with mRMR-based feature selection, discrimination improved. Using the 15-feature MRMR-selected set, the random forest model achieved an AUC of 0.757 (**Figure 2**). Feature importance analysis (**Figure 3**) revealed that model performance was driven by complementary physiologic signals rather than any single dominant variable.

**Figure 1.**
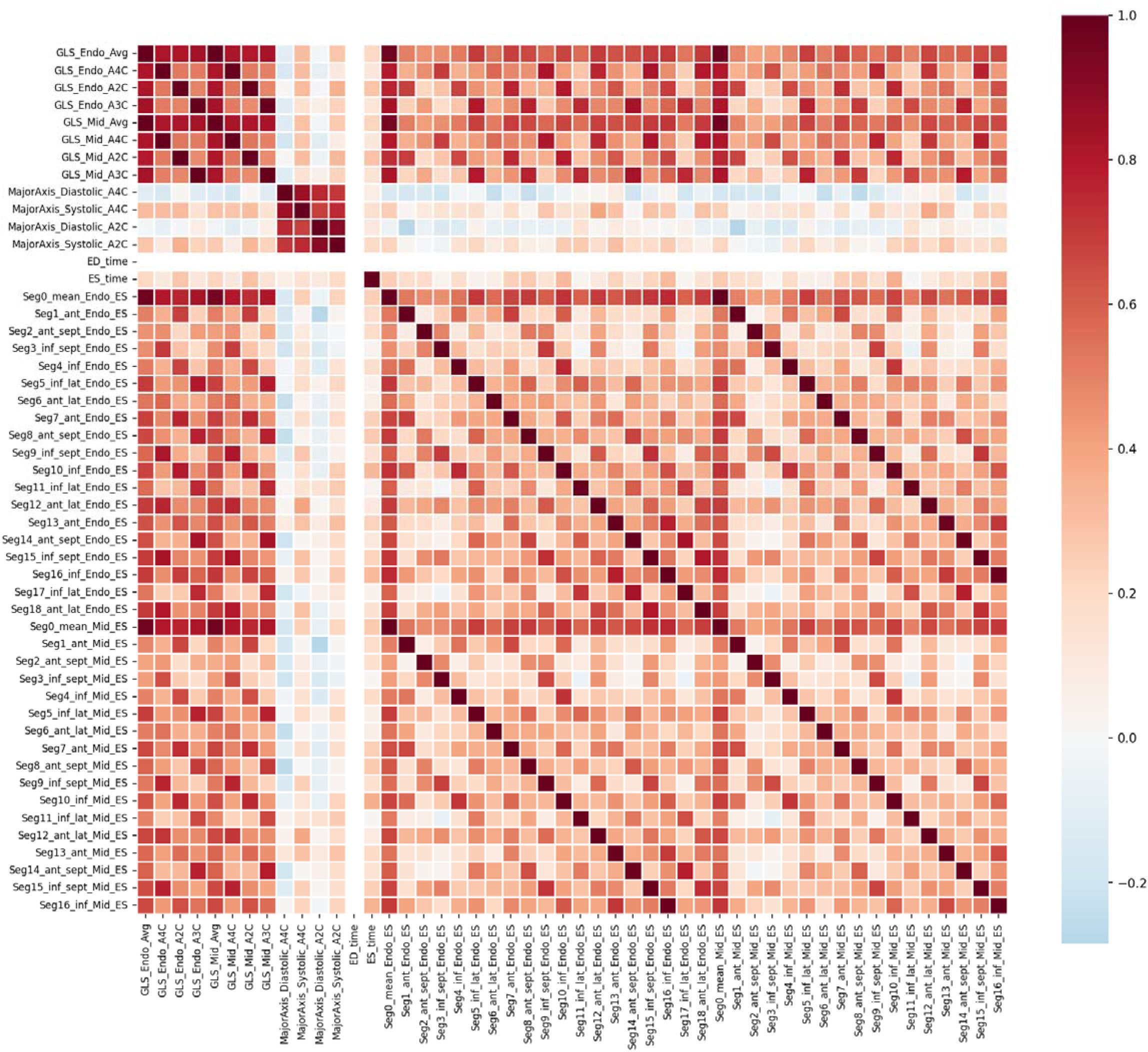
Inter-feature correlation matrix motivating mRMR-based feature selection. Pairwise correlation coefficients among the first 50 echocardiographic strain features are shown, illustrating the high degree of redundancy present in the raw feature space. Clusters of strongly correlated features are evident among segmental end-systolic strain values within the same myocardial layer (endocardial and mid-wall), as well as among global longitudinal strain (GLS) and major axis dimension parameters.

**Figure 2.**
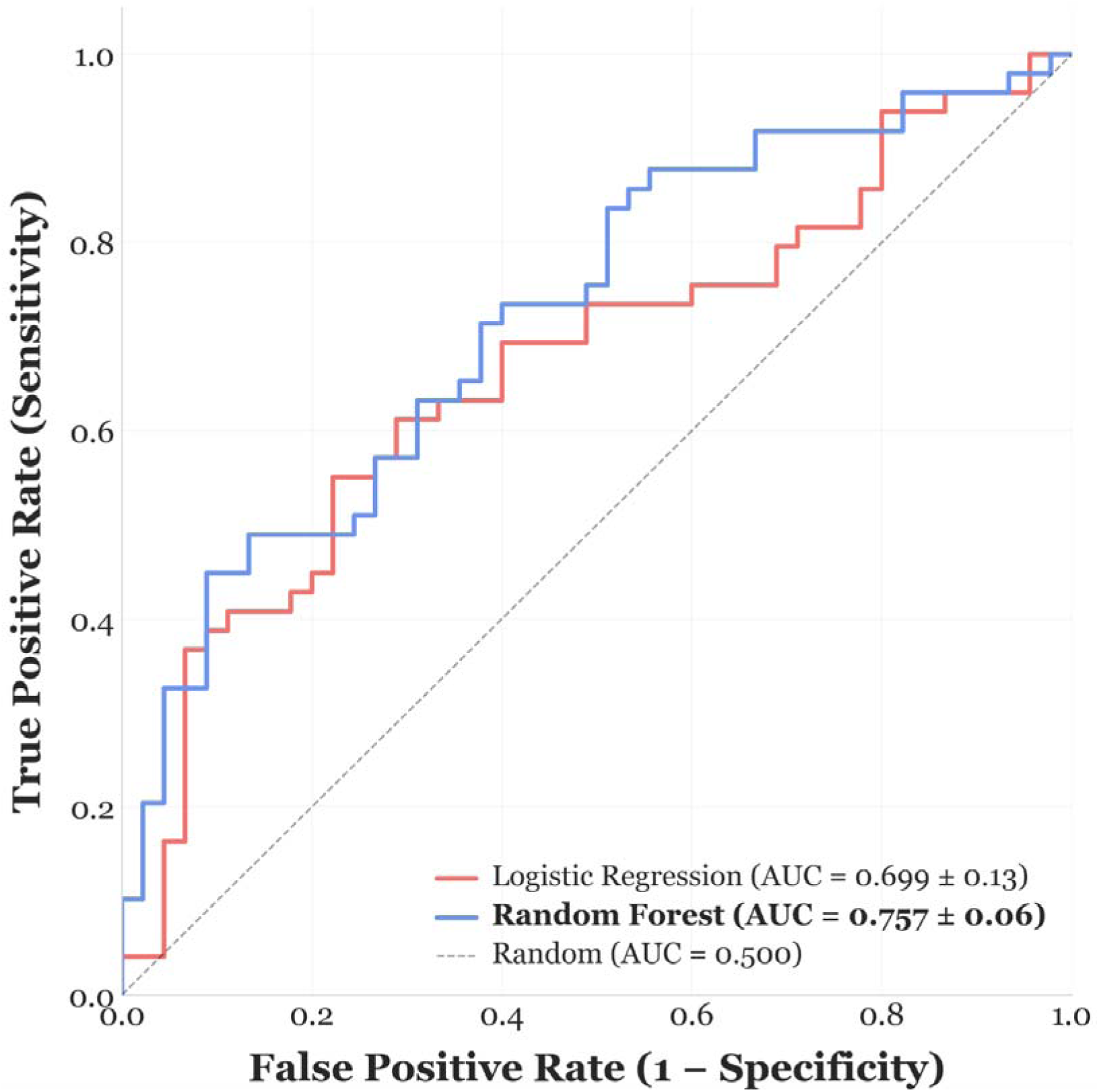
Receiver operating characteristic (ROC) curves for classification of *TTR* status using echocardiographic strain features. ROC curves are shown for random forest (blue) and logistic regression (red) classifiers trained on mRMR-selected strain features and evaluated via leave-one-out cross-validation. The random forest model achieved an area under the curve (AUC) of 0.757 ± 0.06, outperforming logistic regression (AUC = 0.699 ± 0.13). The dashed diagonal line represents chance-level performance (AUC = 0.500).

**Figure 3.**
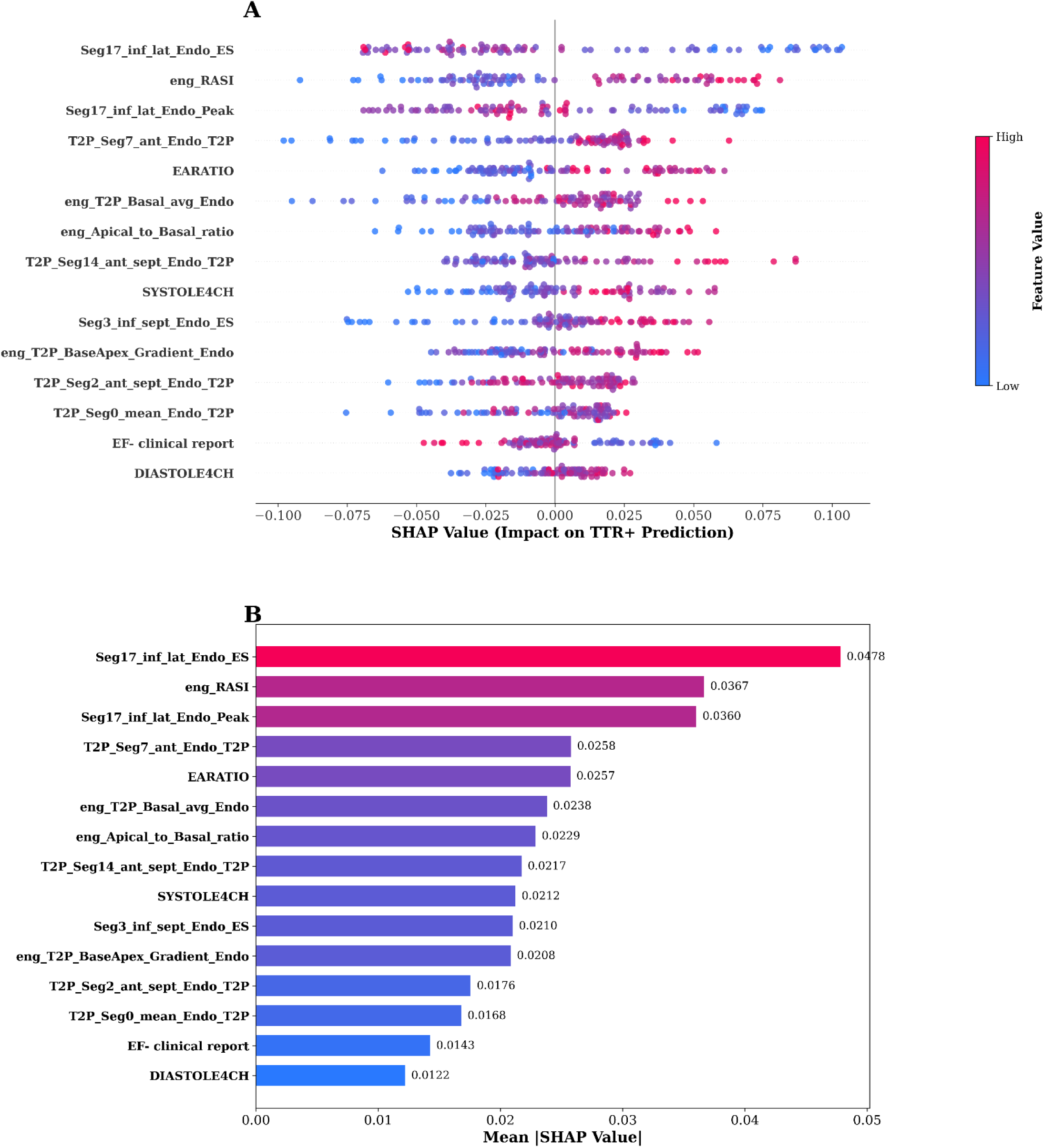
SHAP-based interpretability analysis of the random forest classifier using the top 15 mRMR-selected strain features. **(A)** SHAP beeswarm plot showing the distribution of SHAP values for each feature across all patients. Each dot represents a single patient, with color indicating the feature value (blue = low, magenta = high). Features are ranked by importance from top to bottom. Positive SHAP values indicate contribution toward a *TTR*+ prediction, while negative values indicate contribution toward a *TTR*-prediction. For example, higher values of Seg17_inf_lat_Endo_ES and eng_RASI push predictions toward *TTR*+, whereas higher EARATIO values tend to push predictions toward *TTR*-. **(B)** Mean absolute SHAP value for each feature, quantifying overall feature importance. Seg17_inf_lat_Endo_ES had the highest mean |SHAP| (0.0478), followed by eng_RASI (0.0367) and Seg17_inf_lat_Endo_Peak (0.0360), highlighting the inferolateral wall end-systolic strain and regional strain asymmetry index as the most influential predictors of TTR cardiac amyloidosis.

Prominent contributors included global amyloid signatures such as relative apical sparing and apical-to-basal strain ratios, regional inferolateral strain abnormalities at both end-systole and peak contraction, layer-specific deformation patterns, and mechanical timing features capturing basal-apical gradients in time-to-peak strain. Left ventricular ejection fraction contributed modestly, serving as an anchoring conventional parameter without dominating model predictions.

### Validation

When applied to echocardiographic strain measurements from an external cohort (n=115, *TTR*+ n=59 (84.7% V142I), *TTR*-n=56, 64.3% female, 58.3% AA, 28.7% H/L, average age at time of echo 53.1±12.0 years) the model achieved an AUC=0.781 [95% CI: 0.688-0.869], with a sensitivity of 0.983. (**Figure 4)**. Score stratification did not depend on age (p=0.85), but females trended toward having more low (<0.674) rather than high (≥0.674) risk scores (p=0.07). All non-V142I variant carriers (T80A, n=4; V50M, n=2; F84L, n=1) were classified with high risk scores (V142I vs. non-V142I, p=0.17) (**Table 3**).

**Figure 4.**
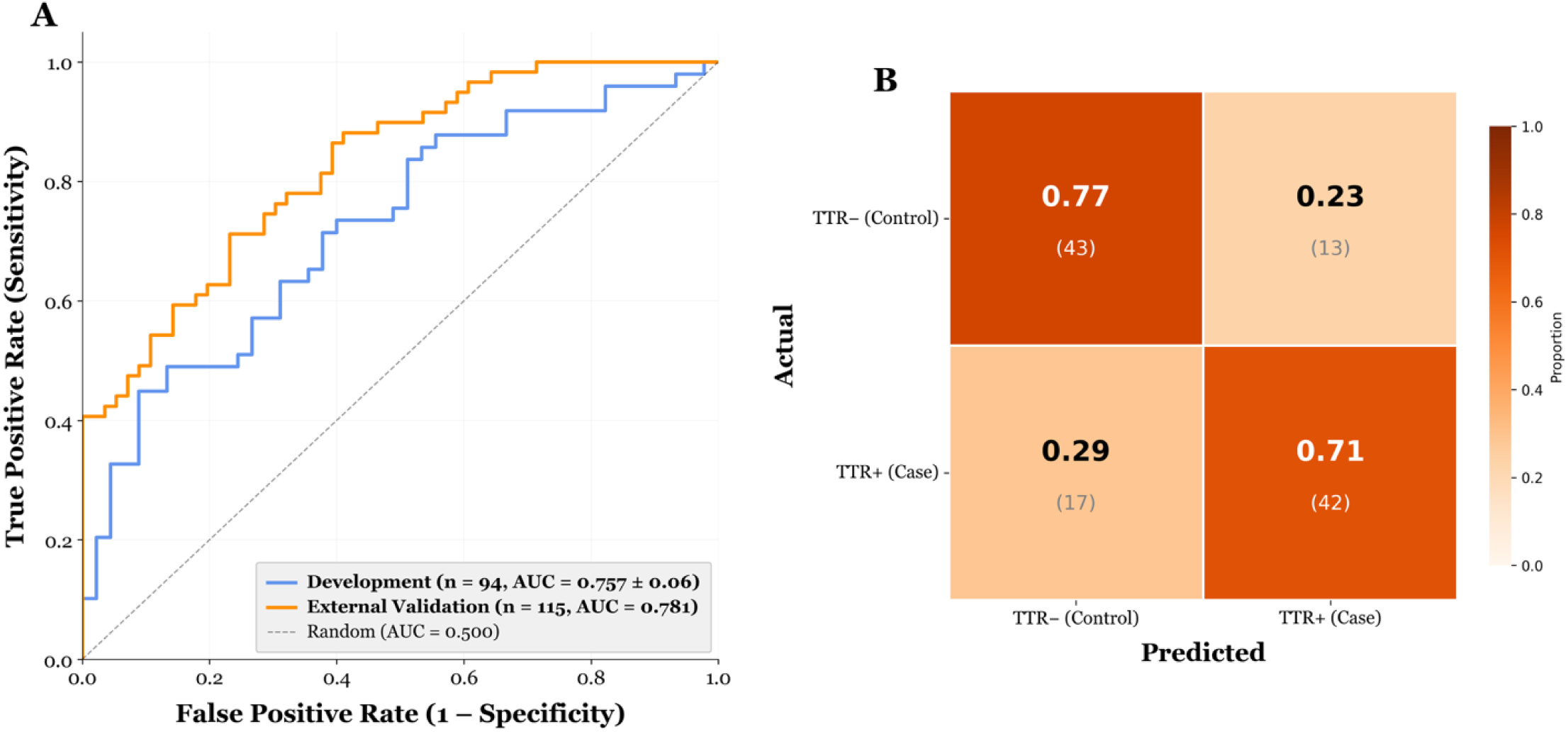
External validation performance of the random forest classifier for *TTR* status. **(A)** Receiver operating characteristic (ROC) curves for the development cohort (n = 94; blue, AUC = 0.757 ± 0.06, nested 5-fold cross-validation) and the independent external validation cohort (n = 115; orange, AUC = 0.781). The dashed diagonal line represents chance-level classification (AUC = 0.500). **(B)** Confusion matrix for the external validation cohort at the optimal operating threshold determined by Youden’s J statistic. Cell values show the proportion of each row correctly or incorrectly classified, with raw counts in parentheses.

**Table 3.**
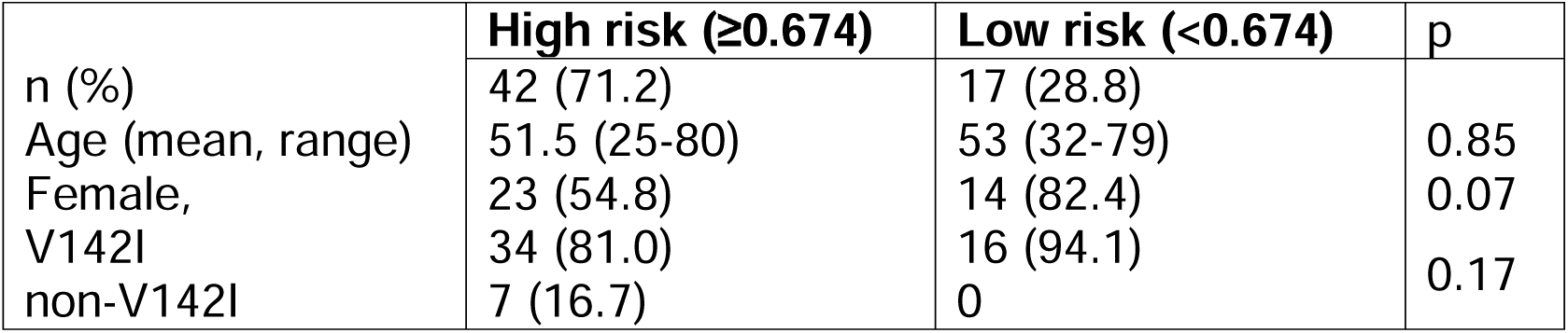
Characteristics of participants in the validation cohort with high vs. low risk scores.

### Longitudinal assessment of the model

Model performance was analyzed in a subset of *TTR*+ carriers with > 1 echocardiogram suitable for STE (n=14, mean per-patient follow-up span 4.9 ±3.0 years, range 1-11 years). Model scores increased over time in all participants (**Figure 5A**). When segregated by age, participants ≥ 55 years old at first available echocardiogram had a greater annual change in their model score compared to participants <55 years old at time of first available echocardiogram (0.0107/year vs. 0.0018/year, p=0.0003, **Figure 5B**).

**Figure 5.**
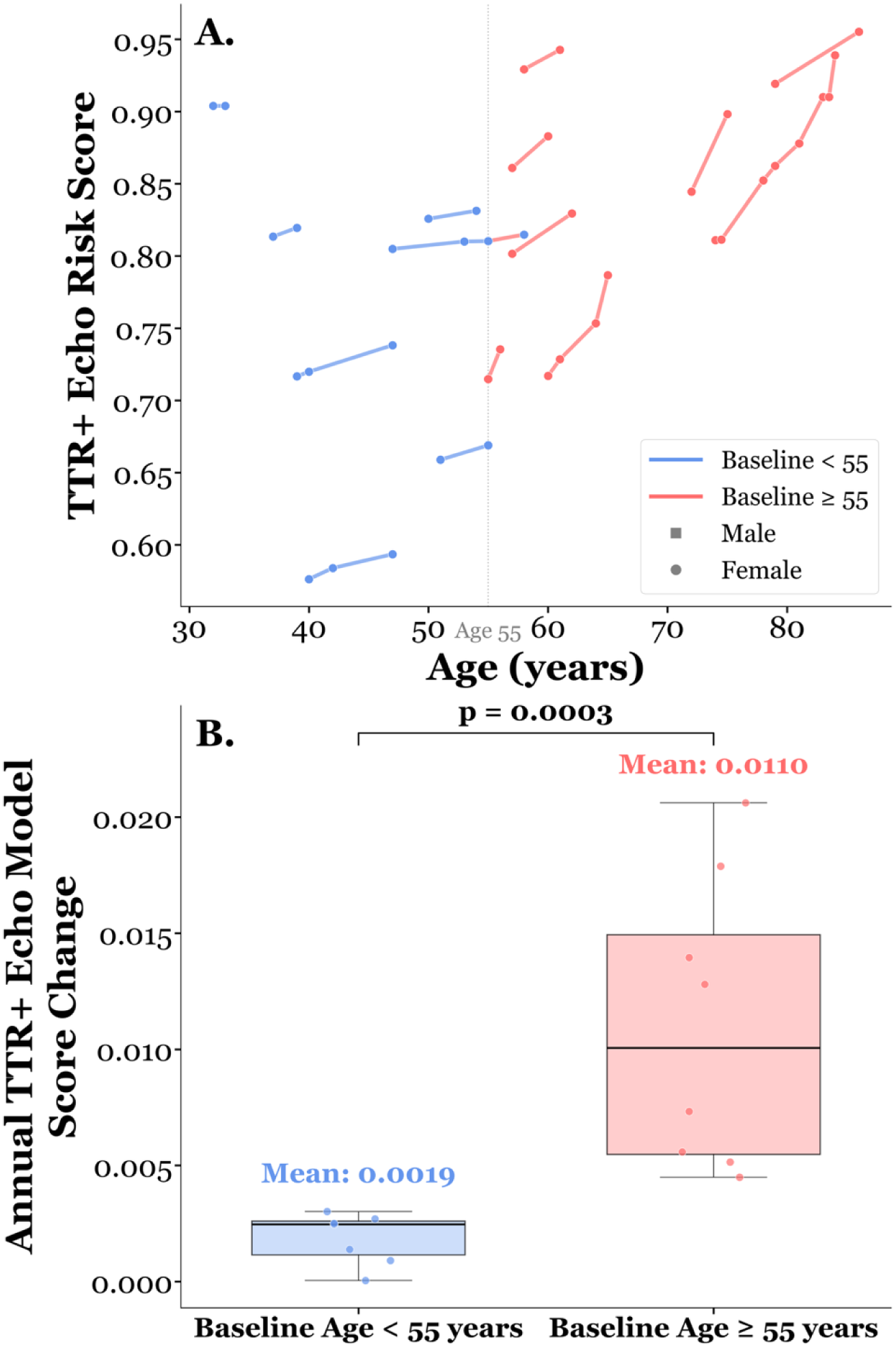
Longitudinal progression of *TTR*+ echo risk scores stratified by baseline age. **(A)** Individual patient trajectories of *TTR*+ echo risk scores plotted against age. Each line connects serial echocardiographic assessments for a single patient, colored by baseline age group (blue: baseline age < 55 years; red: baseline age >= 55 years). Marker shape indicates sex (square = male, circle = female). A vertical reference line at age 55 denotes the stratification threshold. **(B)** Comparison of annual *TTR*+ echo model score change between age groups. Participants with a baseline age >= 55 years demonstrated a significantly higher annual rate of risk score increase (mean = 0.0107/year) compared with those with a baseline age < 55 years (mean = 0.0018/year; p = 0.0003), suggesting accelerated echocardiographic progression toward a ATTR-CA phenotype in older patients, aligning with known age-dependent penetrance of V142I.

## Discussion

Current understanding of ATTRv-related CA is largely derived from traditional “phenotype-first” ascertainment, in which clinical features are characterized in patients with fully manifest disease. Consequently, far less is known about the evolution of signs and symptoms that precede overt CA in high-risk *TTR+* individuals. Better definition of the preclinical phenotype in *TTR*+ carriers is essential to guide optimal surveillance strategies and inform on timing of intervention. “Genotype-first” approaches, which interrogate unselected *TTR*+ individuals for early disease-associated features, can help address this knowledge gap. In our prior evaluation of 337 *TTR* V142I carriers within our large institutional Bio*Me* biobank, HF and cardiomyopathy emerged as the earliest traditional “red flag” manifestations(7), consistent with analyses across Bio*Me* and other biobanks showing that V142I carriers have an approximately 60% lifetime risk of cardiomyopathy and HF(5). Further, young (age 40-60) asymptomatic *TTR* V142I carriers without HF display reduced circulating levels of retinol binding protein 4 (RBP4)(21), an endogenous stabilizer of TTR, suggesting that TTR destabilization is present decades before overt disease develops. Collectively, these observations indicate that the amyloidogenic process begins well before clinical symptom onset, highlighting the need for strategies capable of detecting subtle, early myocardial involvement.

Contemporary diagnostic imaging to evaluate for CA includes echocardiography(22), nuclear scintigraphy or single photon emission computed tomography with bone-avid tracers such as technetium-99 pyrophosphate(23), and cardiac magnetic resonance imaging(24). The utility of these modalities for capturing cardiac changes at early disease stages is undefined, but echocardiography may be particularly well-suited to this application due to the low risk and accessibility of ultrasound technology. In overt disease, echocardiographic features include LV wall thickening, diastolic dysfunction and abnormal GLS and segmental longitudinal strain with preferential basal dysfunction, and relative apical sparing (25,26). These features differentiate CA from other causes of left ventricular hypertrophy and carry prognostic significance(27).

Retrospective registry studies have cited limited cases of asymptomatic non-V142I *TTR*+ individuals in whom echocardiographic abnormalities (LV wall thickening and diastolic impairment) were apparent (28,29). In one small series of asymptomatic *TTR*+ carriers (non-V142I) without LV hypertrophy, GLS was abnormal in more than half of cases(30). However, we previously showed that individual global and regional strain parameters do not reliably distinguish pre-symptomatic *TTR* V142I carriers from matched non-carriers(31). Even qualitative patterns such as apical sparing and quantitative indices such as relative apical sparing index may be present in both groups, underscoring the limited sensitivity of individual strain metrics in early disease. To date, no robust cardiac imaging measures or models have yet been reported to meaningfully discriminate *TTR+* carriers from non-carriers. Strategies focusing on V142I are especially needed, since this is by far the most common cause of ATTRv in the United States.

In this retrospective genotype-first study, we demonstrate that ML applied to high-dimensional echocardiographic strain data can detect subtle myocardial signatures associated with *TTR+* carrier status prior to overt CA. Importantly, this work builds directly on prior negative findings from univariate strain analyses, which showed that global and segmental strain parameters alone are insufficient for early disease detection in pre-symptomatic carriers. The central insight of this study is that early amyloid-related myocardial involvement, when present, is not reflected in isolated abnormalities but instead emerges from distributed, multivariate patterns spanning myocardial regions, layers, and contraction timing. By integrating these features and using mRMR to limit redundancy, we derived a compact, physiologically coherent feature set that enhanced discrimination while reducing overfitting. Several selected features recapitulate known amyloid biology and align with recognized predilection sites of amyloid deposition. For example, relative apical sparing and apical-to-basal strain ratios reflect early spatial heterogeneity as previously described in overt CA (20). Inferolateral strain was also an important feature in our model. Prior work has shown that basal inferolateral segments are among the earliest and most severely affected regions on longitudinal strain analysis in ATTR-CA (32,33), supporting the concept that deformation abnormalities in this region may herald early myocardial involvement. Other features point to potentially novel biological insights. Layer-specific strain metrics suggest differential engagement of endocardial and mid-myocardial fibers, and time-to-peak strain gradients highlight mechanical dyssynchrony as a possible early marker. Notably, conventional ejection fraction contributed only modestly, reinforcing that early disease is not characterized by global systolic dysfunction.

Our model achieved an AUC of 0.76, which should be interpreted in the context of preclinical disease and the known incomplete penetrance of V142I. In this setting, only moderate global discrimination is expected; the goal of ML is risk stratification and prioritization for further evaluation (i.e., genetic testing) rather than definitive diagnosis. This approach parallels emerging genotype-first screening strategies, in which ML enriches the pool of individuals for downstream confirmatory testing rather than replaces clinical judgement(34,35). Importantly, we validated the model in an external *TTR*+ cohort that included carriers of other pathogenic variants (13.6% non-V142I), supporting its generalizability. Moreover, the progressive rise in scores among *TTR*+ carriers with serial echocardiograms suggests that the model is capturing evolving myocardial involvement. These findings align with the known age-dependent penetrance of V142I. Future work will assess implementation of the score in health-system workflows to prioritize patients for *TTR* genomic screening.

Systemic echocardiographic screening for ATTR-CA has been explored previously, but prior efforts have largely relied on late-stage phenotypic features, such as the apical sparing bullseye pattern and interventricular wall thickness ≥ 1.2 cm (36). Recently, another ML model-based approach, ATTRACTnet, was reported. Using tabulated echocardiographic measurements together with discrete diagnosis codes and electrocardiogram waveforms, ATTRACTnet demonstrated good discriminative performance and facilitated new diagnosis of ATTR-CA in 48% of high scoring patients(37). However, both of these strategies are optimized for detecting clinically overt disease and would miss the majority of presymptomatic individuals who have not yet developed significant symptom burden or LV hypertrophy. A model capable of identifying subclinical cardiac manifestations of early ATTR-CA has the potential to enable much earlier recognition of at-risk individuals and ultimately support timely initiation of disease-modifying therapies.

Several limitations of this study should be acknowledged. First, the retrospective design and modest sample size limit causal inference. Additionally, because participants were recruited from a research biobank, their genotype data is generally not incorporated into their clinical records; as a result, most individuals lack ATTRIZIspecific diagnostic evaluations in their EHRs. Consequently, our model cannot directly assess progression to clinical CA. Genotype status—rather than confirmed disease—served as the outcome, reflecting the challenges of studying conditions with incomplete penetrance. Furthermore, echocardiograms were obtained for rather than systematic reasons, introducing potential selection bias. Some, though not all, of these concerns are mitigated by our external validation. Another limitation is the overlap in imaging features across wild-type and variant ATTR, as well as light chain amyloidosis. Thus, myocardial abnormalities observed in *TTR*+ individuals may not be attributable solely to the V142I allele, and unrecognized disease processes may have been present in either *TTR*+ or *TTR*-participants. However, age-, sex- and race/ethnicity matching should reduce differential risk between groups. Ultimately, prospective studies with longitudinal followIZIup will be required to determine whether the imaging patterns identified by the model predict future development of clinical disease.

### Conclusions

ML applied to echocardiographic strain data can identify subtle myocardial abnormalities associated with *TTR*+ carrier status well before overt CA. Echocardiographic strain contains latent information related to genetic risk when analyzed holistically, and mRMR-based feature selection reduces overfitting while grounding the model in established amyloid pathophysiology. Notably, this approach may also reveal novel insights into the natural history and underlying biology of CA beyond current paradigms. Despite the known incomplete penetrance of *TTR* V142I, the model demonstrates robust discriminative performance and uncovers early subclinical phenotypes. As disease-modifying therapies for ATTR continue to expand, ML-enhanced interpretation of routinely acquired imaging may enable scalable, non-invasive surveillance strategies for genetically at-risk populations, supporting genotype-guided monitoring aimed at earlier detection of CA risk.

## Data Availability

The datasets analyzed for the study are not publicly available. The Genomic and EHR data cannot be redistributed to researchers other than those approved through the Icahn School of Medicine at Mount Sinai (ISSMS) Institutional Review Board. We have therefore given detailed description in the manuscript. The code will be made available on GitHub at the time of publication.

## Acknowledgements

We would like to acknowledge the patients who have participated in the Icahn School of Medicine at Mount Sinai Bio*Me* Biobank program.

## Abbreviations list

AA: African American
ATTR: transthyretin amyloidosis
CA: cardiac amyloidosis
GLS: global longitudinal strain
HF: heart failure
H/L: Hispanic/Latino
LV: left ventricular
STE: speckle tracking echocardiography
*TTR*: transthyretin gene

## Notes

**Funding**: This work was supported by NIH grant R01HL155356 and an Investigator Initiated Research award from Akcea/Ionis Therapeutics. The content is solely the responsibility of the authors and does not necessarily represent the official views of the National Institutes of Health.

### Competing Interest Statement

N.S.A.-H. is an employee of the 23andMe Research Institute and a member of the clinical advisory board for Inflection Medicine. A.R.K received research funding from Akcea Therapeutics and Pfizer Inc.

### Funding Statement

This work was supported by NIH grant R01HL155356 and an Investigator Initiated Research award from Akcea/Ionis Therapeutics. The content is solely the responsibility of the authors and does not necessarily represent the official views of the National Institutes of Health.

### Author Declarations

The study was approved by the Mount Sinai Institutional Review Board (GCO 19-01925, GCO 19-00565, ISMMS)

## References

1. Ruberg FL, Grogan M, Hanna M, Kelly JW, Maurer MS. Transthyretin Amyloid Cardiomyopathy: JACC State-of-the-Art Review. J Am Coll Cardiol 2019;73:2872–2891.

2. Rowczenio DM, Noor I, Gillmore JD et al. Online registry for mutations in hereditary amyloidosis including nomenclature recommendations. Hum Mutat 2014;35:E2403–12.

3. Lahuerta Pueyo C, Aibar Arregui MA, Gracia Gutierrez A, Bueno Juana E, Menao Guillen S. Estimating the prevalence of allelic variants in the transthyretin gene by analysing large-scale sequencing data. Eur J Hum Genet 2019;27:783–791.

4. Yamashita T, Hamidi Asl K, Yazaki M, Benson MD. A prospective evaluation of the transthyretin Ile122 allele frequency in an African-American population. Amyloid 2005;12:127–30.

5. Damrauer SM, Chaudhary K, Cho JH et al. Association of the V122I Hereditary Transthyretin Amyloidosis Genetic Variant With Heart Failure Among Individuals of African or Hispanic/Latino Ancestry. JAMA 2019;322:2191–2202.

6. Parker MM, Damrauer SM, Tcheandjieu C et al. Association of the transthyretin variant V122I with polyneuropathy among individuals of African ancestry. Sci Rep 2021;11:11645.

7. Kontorovich AR, Benson CB, McClellan A, Belbin GM, Kenny EE, Abul-Husn NS. Evolving knowledge of “red flag” clinical features associated with TTR p.(Val142Ile) in a diverse electronic health record-linked biobank. Genet Med 2024:101346.

8. Grodin JL, Gupta A, Rison I, Jr. et al. Risk for Heart Failure and Atrial Fibrillation Across the Lifespan for Carriers of the Amyloidogenic p.V142I TTR Variant. Circ Genom Precis Med 2025;18:e004911.

9. Selvaraj S, Claggett B, Shah SH et al. Cardiovascular Burden of the V142I Transthyretin Variant. JAMA 2024;331:1824–1833.

10. Dorbala S, Ando Y, Bokhari S et al. ASNC/AHA/ASE/EANM/HFSA/ISA/SCMR/SNMMI Expert Consensus Recommendations for Multimodality Imaging in Cardiac Amyloidosis: Part 1 of 2-Evidence Base and Standardized Methods of Imaging. Circ Cardiovasc Imaging 2021;14:e000029.

11. Maurer MS, Sultan MB, Rapezzi C. Tafamidis for Transthyretin Amyloid Cardiomyopathy. N Engl J Med 2019;380:196–197.

12. Gillmore JD, Judge DP, Cappelli F et al. Efficacy and Safety of Acoramidis in Transthyretin Amyloid Cardiomyopathy. N Engl J Med 2024;390:132–142.

13. Fontana M, Berk JL, Gillmore JD et al. Vutrisiran in Patients with Transthyretin Amyloidosis with Cardiomyopathy. N Engl J Med 2025;392:33–44.

14. Maurer MS, Kale P, Fontana M et al. Patisiran Treatment in Patients with Transthyretin Cardiac Amyloidosis. N Engl J Med 2023;389:1553–1565.

15. Maurer MS, Schwartz JH, Gundapaneni B et al. Tafamidis Treatment for Patients with Transthyretin Amyloid Cardiomyopathy. N Engl J Med 2018;379:1007–1016.

16. Fontana M, Berk JL, Gillmore JD et al. Vutrisiran in Patients with Transthyretin Amyloidosis with Cardiomyopathy. N Engl J Med 2024.

17. Elliott P, Drachman BM, Gottlieb SS et al. Long-Term Survival With Tafamidis in Patients With Transthyretin Amyloid Cardiomyopathy. Circ Heart Fail 2022;15:e008193.

18. Maurer MS, Bokhari S, Damy T et al. Expert Consensus Recommendations for the Suspicion and Diagnosis of Transthyretin Cardiac Amyloidosis. Circ Heart Fail 2019;12:e006075.

19. Falk RH. Tafamidis for transthyretin amyloid cardiomyopathy: the solution or just the beginning of the end? Eur Heart J 2019;40:1009–1012.

20. Phelan D, Collier P, Thavendiranathan P et al. Relative apical sparing of longitudinal strain using two-dimensional speckle-tracking echocardiography is both sensitive and specific for the diagnosis of cardiac amyloidosis. Heart 2012;98:1442–8.

21. Kontorovich AR, Abul-Husn NS. Retinol Binding Protein 4 as a Screening Biomarker for Hereditary TTR Amyloidosis in African American Adults With TTR V142I. J Card Fail 2021;27:1020–1022.

22. Falk RH, Quarta CC. Echocardiography in cardiac amyloidosis. Heart Fail Rev 2015;20:125–31.

23. Gillmore JD, Maurer MS, Falk RH et al. Nonbiopsy Diagnosis of Cardiac Transthyretin Amyloidosis. Circulation 2016;133:2404–12.

24. Zhao L, Tian Z, Fang Q. Diagnostic accuracy of cardiovascular magnetic resonance for patients with suspected cardiac amyloidosis: a systematic review and meta-analysis. BMC Cardiovasc Disord 2016;16:129.

25. Bellavia D, Abraham TP, Pellikka PA et al. Detection of left ventricular systolic dysfunction in cardiac amyloidosis with strain rate echocardiography. J Am Soc Echocardiogr 2007;20:1194–202.

26. Ternacle J, Bodez D, Guellich A et al. Causes and Consequences of Longitudinal LV Dysfunction Assessed by 2D Strain Echocardiography in Cardiac Amyloidosis. JACC Cardiovasc Imaging 2016;9:126–38.

27. Pagourelias ED, Mirea O, Duchenne J et al. Echo Parameters for Differential Diagnosis in Cardiac Amyloidosis: A Head-to-Head Comparison of Deformation and Nondeformation Parameters. Circ Cardiovasc Imaging 2017;10:e005588.

28. Mazzeo A, Russo M, Di Bella G et al. Transthyretin-Related Familial Amyloid Polyneuropathy (TTR-FAP): A Single-Center Experience in Sicily, an Italian Endemic Area. J Neuromuscul Dis 2015;2:S39–S48.

29. Minutoli F, Di Bella G, Mazzeo A et al. Serial scanning with (99m)Tc-3, 3-diphosphono-1, 2-propanodicarboxylic acid ((99m)Tc-DPD) for early detection of cardiac amyloid deposition and prediction of clinical worsening in subjects carrying a transthyretin gene mutation. J Nucl Cardiol 2019.

30. Canciello G, Tozza S, Todde G et al. Global longitudinal strain in pre-symptomatic patients with mutation for transthyretin amyloidosis. Orphanet J Rare Dis 2024;19:458.

31. Kontorovich AR, Zhao W, Gavalas M et al. Left ventricular strain does not differentiate amyloidogenic profiles in at-risk individuals with TTR Val142Ile. medRxiv 2021:2021.01.26.21250565.

32. Usuku H, Yamamoto, E., Imamura, K., Higashi, R., Nozuhara, A., Oike, F., Kuyama, N., Ishii, M., Hanatami, S., Hoshiyama, T., Kanazawa, H., Arima, Y., Oda, S., Kawano, H., Matsuzawa, Y., Izumiya, Y., Ueda, M., Tanaka, Y., Tsujita, K. Usefulness of Regional Longitudinal Strain to Identify Transthyretin Amyloid Cardiomyopathy. Circulation Reports 2025;CR-25-0269.

33. Tsuruda T, Nakada H, Yamamura Y et al. Basal inferoseptal segment is highly susceptible to deformation in the clinical spectrum of transthyretin-derived amyloid cardiomyopathy. Eur Heart J Open 2024;4:oeae076.

34. Attia ZI, Noseworthy PA, Lopez-Jimenez F et al. An artificial intelligence-enabled ECG algorithm for the identification of patients with atrial fibrillation during sinus rhythm: a retrospective analysis of outcome prediction. Lancet 2019;394:861–867.

35. Grogan M, Lopez-Jimenez F, Cohen-Shelly M et al. Artificial Intelligence-Enhanced Electrocardiogram for the Early Detection of Cardiac Amyloidosis. Mayo Clin Proc 2021;96:2768–2778.

36. Agarwal A, Jia K, Vachss D et al. Systematic Echocardiographic Screening Protocol for Cardiac Amyloidosis Detection: A Low-Cost, High-Impact Innovation. JACC Case Rep 2025;30:105657.

37. Jain SS, Sun T, Pierson E et al. Detecting Transthyretin Cardiac Amyloidosis With Artificial Intelligence: A Nonrandomized Clinical Trial. JAMA Cardiol 2026;11:117–124.

